# The Invisible Wounds: Prevalence, Determinants, and Lived Experiences of Post-Traumatic Stress Disorder in Bahir Dar, Ethiopia: A Mixed-Methods Study

**DOI:** 10.1101/2025.08.23.25333557

**Authors:** Medhanit Tadesse, Melak Menberu Guangu, Minale Tareke

## Abstract

**Background:** Post traumatic stress disorder (PTSD) develops due to exposure to a traumatic experience, leading to functional impairment and diminishes the quality of life. Most previous studies were not conducted during ongoing armed conflict and do not provide a detailed exploration of trauma survivors’ actual experiences. As a result, it is critical to understand the prevalence of PTSD and its impact on the lives of Bahir Dar residents.

**Methods:** A convergent mixed study design was conducted in Bahir Dar city Northwest Ethiopia. The participants were selected from the general population. The PTSD Checklist for DSM-5 (PCL-5) was utilized to assess the PTSD and in-depth interviews were conducted to explore the lived experiences of the participants. Logistic regression analysis was used to identify determinant factors, and all significance tests with a p-value < 0.05 were considered significant factors associated with PTSD.

**Results:** The prevalence of PTSD was 36.2%. Being female divorced/widowed marital status, depression, suicidal behavior, stressful life event, poor social support, perceived stress, and problematic substance use were statistically significantly associated with PTSD. The in-depth interview identified four themes: traumatic experiences, PTSD symptoms, coping mechanisms, and impact on life.

**Conclusion:** the prevalence of PTSD was found to be high among residents in Bahir Dar city. Therefore, it is essential to develop comprehensive support systems that address these mental health challenges and promote recovery.

## Introduction

Post-traumatic stress disorder is a mental health issue stemming from exposure to distressing events such as violent assaults with war and conflict being particularly main contributors (1, 2). The symptoms of PTSD include intrusion, avoidance, mood fluctuations, cognitive disruptions, and heightened arousal, persisting for over a month after the traumatic incident.

Globally, PTSD accounts for a substantial portion of the mental health disease burden, affecting approximately 3.9% of the general population, but with much higher rates in post-conflict areas and among those directly exposed to traumatic events (3, 4). In Sub-Saharan Africa, and especially in war affected regions, prevalence can reach up to 30% (5). The impact of PTSD is profound, resulting in diminished quality of life, increased reliance on medical and social support, and impaired work performance. Additionally, PTSD is associated with significant biological, psychological, behavioral, and social repercussions, leading to considerable functional limitations (6, 7).

The factors that mainly contributes for the development of PTSD, includes genetic, environmental, and other variables such as the nature of traumatic exposure, poor self-management, socio cultural and physical disabilities. Genetic and environmental factors significantly impact PTSD (8).

There is a growing burden of PTSD in many countries experiencing an armed conflict and one of them is Ethiopia. However, research on the prevalence, associated factors, and lived experiences of individuals with PTSD in Ethiopian conflict zones remains limited. Existing studies offer only fragmented prevalence estimates and rarely explore the contextual and experiential dimensions of trauma in these settings. This gap in knowledge hinders the development of effective, context specific mental health interventions.

This study addresses these gaps by investigating the prevalence and determinants of PTSD among residents of Bahir Dar, Ethiopia a city directly affected by ongoing conflict. By combining quantitative analysis with qualitative exploration of individuals’ lived experiences, this research aims to provide a comprehensive understanding of PTSD in this context.

The findings of this study can help guide decisions on policy that support and rehabilitate populations harmed by conflict. It would also direct the distribution of funds for projects that promote psycho-social assistance, mental health care, and peace building.

## Materials and Methods

### Study design, setting and period

A convergent mixed methods study was conducted from October 1 to 20, 2024 in Bahir Dar city, Ethiopia. The study combined quantitative data collected through household surveys with qualitative data obtained from in depth interviews to provide comprehensive insights.

### Target population

The study population included all permanent residents of Bahir Dar city who had lived there for at least six months. For the quantitative component, randomly selected households served as sampling units, with individuals aged 18 years and older within these households comprising the study units. For the qualitative component, purposively selected individuals provided in-depth perspectives.

### Inclusion and exclusion criteria

Eligible participants included permanent residents of Bahir Dar city for at least six months and individuals aged 18 years or older. We excluded participants who were severely ill or otherwise unable to communicate effectively, as they could not provide reliable information for the study.

### Sampling Procedure

For the quantitative part the study, participants were recruited using a multi-stage sampling process. Initially, two out of six sub-cities in Bahir Dar City were chosen using simple random sampling. Kebeles inside these identified sub-cities were then also chosen using simple random sampling. The sample size was proportionally distributed to households in the chosen, and households were selected using systematic random selection from a sampling frame received from the city municipal and local health extension office. If there were more than two eligible persons in a given home, one was picked by lottery method. Interviewers attempted up to three revisits for unavailable participants before declaring a home non-responsive. The qualitative component was carried out using a purposive sampling method. Participants with a PCL-5 score of 33 or above, suggesting significant PTSD symptoms were identified from the quantitative sample and asked for in-depth interviews to discuss their trauma and lived experiences of armed conflict. Data saturation was used to establish the qualitative sampling size.

**Figure 1:**
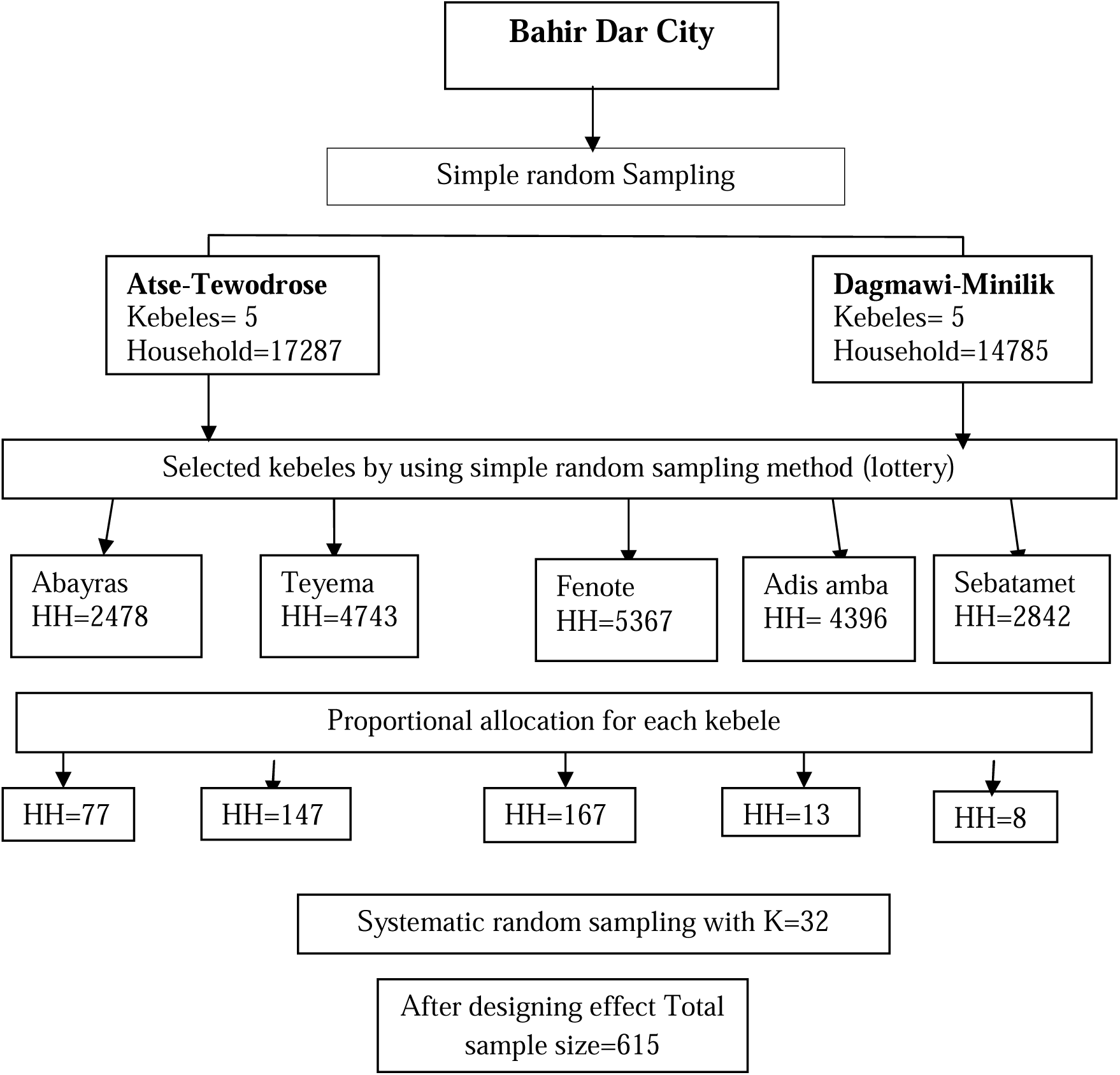
Schematic presentation of sampling procedure on the prevalence of PTSD and associated factors among residents in Bahir Dar, Ethiopia, 2024.

### Data Collection Tools and Procedures

Data was collected by face-to-face interviews using structured and semi-structured questionnaire by four BSc psychiatric nurses and was supervised by two MSc psychiatric professionals.

PTSD was measured using the PCL-5 post-traumatic stress disorder checklist. PCL5 is a standardized self-assessment tool and scale for assessing 20 symptoms of PTSD. The total score is calculated by summing 20 items, and the possible scores range from 0 to 80 on the Likert scale of 5 points (0 = Not at all, 1 = A little bit, 2 = moderately, 3 = Quite a bit, 4 = extremely) with a cutoff point of ≥33 with sensitivity of 74.5% and a specificity of 70.6% (9). Validity and reliability of the PCL-5 had been tested and proven on displaced people and refugees in a number of countries and this tool was used on the study done in Ethiopia (1, 10). Reliability coefficient Cronbach s α in this study for the DSM5 (PCL5) was 0.91 showing a good internal consistency of the items.

Depression was measured by PHQ-9, which indicates depression if individuals scoring at cut point 10 or more out of 9 items. It has 4 items consisted of Not at all (0), Several Days (1), More than half the days, (2) Nearly every day (3) over the last 2 weeks. It has been validated in Ethiopia to screen for diagnosing major depressive disorder among adults in Ethiopia (11). Reliability coefficient Cronbach s α in this study PHQ was 0.83 showing a good internal consistency of the items.

Suicidal behavior among population was assessed by Suicide Behaviors Questionnaire-Revised (SBQ-R) tool with Cronbach’s alpha score of 0.88 in the Ethiopian validation study, indicating a high level of internal consistency reliability. The sensitivity and specificity of the SBQR tool in the Ethiopian validation study were reported 86% and 92% respectively (12).

Social support was measured using the Oslo3 social support scale, which ranges from 3 to 14. Respondents with a score of 3–8 were considered to have poor social support, scores 9–11 were considered moderate social support, and scores 12–14 are considered strong social support (13).

To examine substance use history, respondents was asked: “Have you ever used any substance in the last three months or in your lifetime?” and the responses was yes or no. In addition, CAGE-AID:(Cut down, Annoyed, Guilty, and Eye-opener) questionnaire used to screen for problematic substance uses and other problematic drug uses (khat, tobacco and cannabis) in addition to screen problematic alcohol uses. Item responses on the CAGE questions were scored 0 for “no” and 1 17 for “yes” answers. A total score of two or greater positive answers from the four questions to social drugs (khat, alcohol, tobacco and cannabis) were considered as problematic substance uses (14, 15).

List of Threatening Experiences (LTE) questionnaire was used to assess stressful events with yes/no answers of respondents. The LTE consisted of 12 individual stressful events items (16). It is a valid and reliable measure of stress in mental health (62), and it has been utilized in Ethiopia (17). Reliability coefficient Cronbach s α in this study for the LTE was 0.81 showing a good internal consistency of the items.

An individual’s stress level was measured using the Perceived Stress Scale (PSS-10). The questions on this scale asked about feelings and thoughts of last month. PSS was measured with a Likert type scale ranging from (0) “never” to (4) “very often”. The four positively stated items (item 4, 5, 7, and 8) were reversed (e.g., 0 = 4, 1 = 3, 2 = 2, 3 = 1, and 4 = 0) and then summed across all scale items. Individuals with a score of “0-13” were considered to have low perceived stress, “14-26” moderate perceived stress, and “27-40” higher perceived stress (18).

Data Collection Tools and Procedure for Qualitative study

Semi-structured interviews were served as the primary data collection tool using epicollect5, meticulously designed to capture detailed personal insights into individuals’ experiences with PTSD. The interview guide was designed to explore various facets of PTSD, such as symptoms, triggers, coping mechanisms, and their impacts on daily life.

The data collection procedure involves several structured steps to effectively utilize this tool. It started with the development of the interview guide, which ensures that questions were specific and relevant. Participants were selected through purposive sampling to include a diverse range of experiences and perspectives. Interviews were held in a quiet and friendly environment. Each interview was recorded with the participants’ agreement to ensure data accuracy while adhering to rigorous confidentiality and data protection rules.

### Data handling and analysis

Data were collected by trained psychiatric nurses and supervised by psychiatric professionals. The questionnaire was translated into Amharic and back-translated to English for consistency. Data were checked for completeness, coded, and entered into Epi-data version 4.6, then exported to SPSS version 25 for analysis. Descriptive statistics summarized study variables. Variables with p ≤ 0.25 in bivariable logistic regression were included in multivariable analysis to identify factors associated with PTSD, with significance set at p < 0.05. Model fit was confirmed by the Hosmer-Lemeshow test (χ² = 4.574, p = 0.802), and no multicollinearity was detected (tolerance > 0.10, VIF 1.06–1.43). Qualitative data were transcribed verbatim and analyzed using ATLAS.ti9. Thematic analysis involved coding, theme development, and validation through member checking and triangulation to ensure credibility.

### Ethical Consideration

The study was commenced after obtaining ethical clearance from the Institutional Review Board (IRB) of Bahir Dar University College of Medicine and Health Science (IRB Protocol number: 2074/2024). An ethical clearance letter was submitted to the Bahir Dar city health administration for permission. The study’s objectives were explained to the administrative body, and a supportive letter from them was disseminated to the study areas. Participants were individually approached, briefed on the study’s purpose and the confidentiality of their information, and asked for consent. Participants were informed that they would not get any benefit because of participation in the study and no harm on them if they do not agree to participate or withdraw from participation during the data collection. When the participants’ data were finished, the epicollect5 alert requested that data collectors link to the nearby health facility if the person is experiencing severe depression or suicidal thoughts.

## Results

### 5.1. Quantitative Data Results

#### 5.1.1. Socio-demographic characteristics of the participants

A total of 585 participants were involved with response rate of 95.1%. The mean age of the respondents was 37.15 years with SD ± 11.93 years, and the nearly one third of the respondents 214 (36.6%) were between the age of 26-35 years. More than half of the respondents were male 329 (56.2%), also large majority of the participants were married 372(63.6%), having diploma and above in their educational status were 220(37.6%), and nearly half of the respondents were employed 284 (48.5%), (Table 2)

**Table 1:**
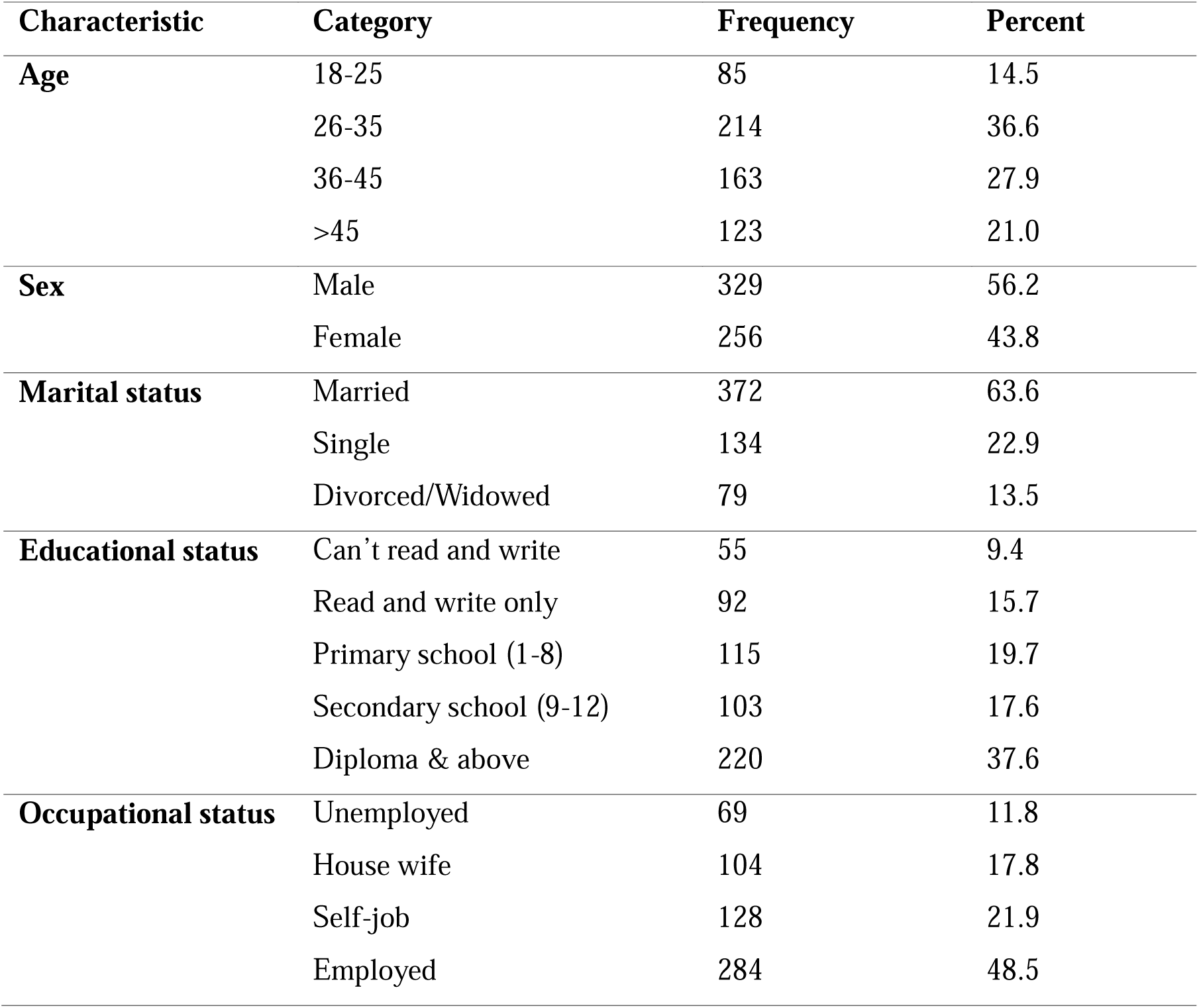
Socio-demographic characteristics of study participants among residents in Bahirdar city, Northwest, Ethiopia, 2024 (n=585).

**Table 2:** Clinical and substance related factors of the respondents of the residents in Bahir Dar city, Northwest, Ethiopia 2024 (n=585).

| Characteristic | Category | Frequency | Percent |
| --- | --- | --- | --- |
| Family history of diagnosed known mental illness | Yes | 71 | 12.1 |
|  | No | 514 | 87.9 |
| Known diagnosed chronic medical condition | Yes | 76 | 13.0 |
|  | No | 509 | 87.0 |
| Depression | Yes | 195 | 33.3 |
|  | No | 390 | 66.7 |
| Suicidal behavior | Yes | 103 | 17.6 |
|  | No | 482 | 82.4 |
| Alcohol drinks in lifetime | Yes | 369 | 63.1 |
|  | No | 216 | 36.9 |
| Current alcohol use | Yes | 344 | 58.8 |
|  | No | 241 | 41.2 |
| Problematic substance use disorder | Yes | 97 | 16.6 |
|  | No | 488 | 83.4 |

##### Clinical and substance related factors of the respondents

Approximately one-third (33.3%) of the respondents indicated experiencing symptoms consistent with depression, while near to one-fifth (17.6%) respondents expressed suicidal behavior (table 3).

**Table 3:** Psychosocial factors of the respondents in Bahirdar city, Northwest, Ethiopia 2024 (n=585).

| Characteristic | Category | Frequency | Percent |
| --- | --- | --- | --- |
| Stressful life event | Yes | 311 | 53.2 |
|  | No | 274 | 46.8 |
| Social support | Poor | 200 | 34.2 |
|  | Moderate | 231 | 39.5 |
|  | Strong | 154 | 26.3 |

##### Psychosocial factors of PTSD

From the participants more than half of the participants 311(53.2%) had experienced at least one stressful life event during the last 6 months and about 234(40%) of the respondents have poor social support.

**Figure 2:**
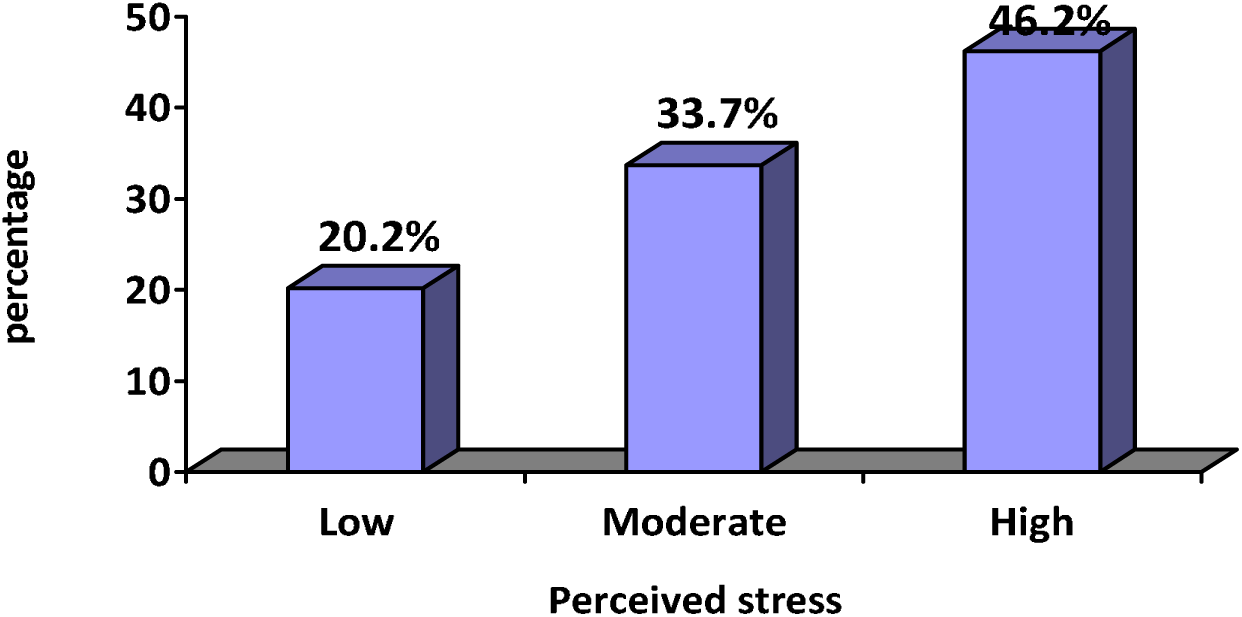
Description of the perceived stress of respondents in Bahirdar city, Northwest, Ethiopia, 2024 (n=585)

### 5.1.6. Prevalence of PTSD

The prevalence of PTSD was **36.2%** (CI: 32.2%-40.0%). Being Female, Divorced/widowed, depression, suicidal behavior, stressful life event, poor social support, perceived stress, and problematic substance use were statistically significantly associated with PTSD.

Bi-variable and multivariable binary logistic regression analysis showing association between independent factors and PTSD among residents living in Bahir Dar city, Ethiopia 2024 (n=585)

| Variable | Category | PTSD |  | COR (95%CI) | AOR (95%CI) | P value |
| --- | --- | --- | --- | --- | --- | --- |
|  |  | Yes | No |  |  |  |
| Sex | Female | 116 | 140 | 2.01(1.43,2.83) | 1.91(1.18,3.08) | 0.008 |
|  | Male | 96 | 233 | 1 | 1 |  |
| Marital status | Single | 47 | 87 | 1.13(0.58,1.33) | 1.16(0.70,1.90) | 0.06 |
|  | Divorced/<br>widowed | 45 | 34 | 2.77(1.69,4.56) | 2.03(1.13,3.63) | 0.01 |
|  | Married | 120 | 252 | 1 | 1 |  |
| Family Hx of<br>diagnosed MI | Yes | 37 | 34 | 2.10(1.27,3.47) | 1.26(0.64,2.49) | 0.49 |
|  | No | 175 | 339 | 1 | 1 |  |
| Depression | Yes | 111 | 84 | 3.78(2.63,5.43) | 1.79(1.15,2.79) | 0.01 |
|  | No | 101 | 289 | 1 | 1 |  |
| Suicidal behavior | Yes | 64 | 39 | 3.70(2.37,5.76) | 1.95(1.08,3.51) | 0.02 |
|  | No | 148 | 334 | 1 | 1 |  |
| Stressful life event | Yes | 143 | 168 | 2.52(1.77,3.59) | 2.17(1.39,3.40) | <0.01 |
|  | No | 69 | 205 | 1 | 1 |  |
| Social support | Poor | 109 | 91 | 6.18(3.70,10.30) | 3.36(1.70,6.66) | <0.001 |
|  | Intermediate | 78 | 153 | 2.35(1.9,3.46) | 1.16(0.66,1.87) | 0.68 |
|  | Strong | 25 | 129 | 1 | 1 |  |
| Perceived stress scale | High | 146 | 124 | 6.54(3.75,11.40) | 2.91(1.70,4.96) | <0.001 |
|  | Moderate | 48 | 149 | 1.78(1.44,2.47) | 1.98(0.96,2.08) | 0.06 |
|  | Low | 18 | 100 | 1 | 1 |  |
| Problematic substance use | Yes | 57 | 40 | 3.06(1.95,4.78) | 2.24(1.26,3.97) | 0.006 |
|  | No | 155 | 333 | 1 | 1 |  |

#### Qualitative data results

##### Description of participants

Data were collected from six participants who had symptoms of PTSD with a score of >33 on PCL 5, comprising four males and two females, with ages ranging from 20’s-50’s. Their socio demographic characteristics were presented in table.

**Table 4:**
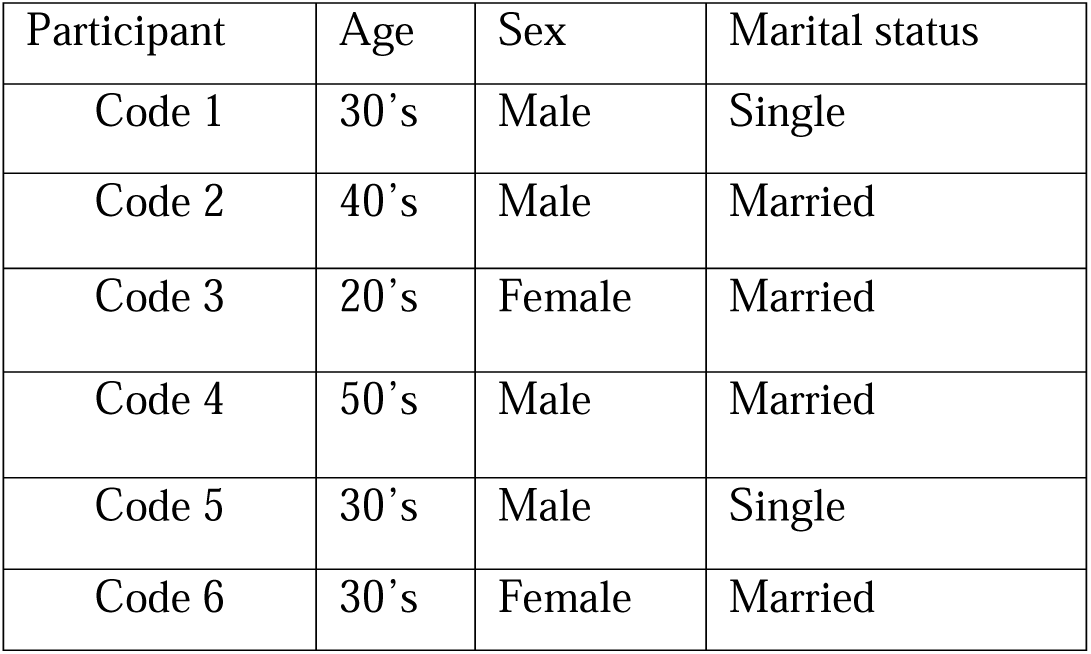
Socio – demographic characteristics of the participants (qualitative data)

| Participant | Age | Sex | Marital status |
| --- | --- | --- | --- |
| Code 1 | 30's | Male | Single |
| Code 2 | 40's | Male | Married |
| Code 3 | 20's | Female | Married |
| Code 4 | 50's | Male | Married |
| Code 5 | 30's | Male | Single |
| Code 6 | 30's | Female | Married |

##### Themes and Subthemes identified

**Table 5:** Themes and sub themes identified from in-depth interview.

| Theme | Description | Sub-themes |
| --- | --- | --- |
| **1. Traumatic Experiences** | Participants recount specific traumatic events, detailing when and where they occurred, and personal accounts of what happened, often highlighting feelings of shock and fear during the event. | • Description of the<br>• Traumatic Event<br>• Immediate Reactions |
| **2. PTSD Symptoms** | PTSD Symptoms encompass a range of psychological responses that arise following traumatic events. The emotional toll includes heightened anxiety, irritability, and depression, which can strain relationships and hinder daily activities. This theme illustrates the ongoing battle individuals face as they cope with the lasting effects of trauma on their mental health. | • Intrusive Thoughts and Memories<br>• Emotional Responses and mood change<br>• Sleep difficulties |
| **3. Coping Mechanisms** | This Theme explores the various strategies individuals use to manage the effects of trauma on their lives. it underscores the complexity of recovery, highlighting the importance of finding effective coping methods while navigating the challenges posed by trauma. | • Support system<br>• Active coping strategies<br>• Avoid triggers<br>• Substance use |
| **4. Impact on Life** | This theme emphasizes the extensive consequences of PTSD on personal relationships and overall quality of life, underscoring the need for understanding and support during recovery. | • Relationships<br>• Work and Daily Functioning |

###### 1. Traumatic Experiences

Participants described their traumatic experiences that led to post traumatic stress symptoms. They frequently mentioned a sudden, explosive event that completely disrupted their daily lives. The participants reported traumatic visual encounter with death during the attack and also stated witnessing violence and the fear of being targeted by gunfire. One individual recounted their experience, remarking:

> *“We saw people being shot, and everyone was running to hide from the gunfire…“We looked out and saw many bodies lying on the ground; … it was a horrifying sight.” Male, (50’s)*

Participants reported immediate reactions that include feelings of shock, fear, and instinctive actions taken to ensure safety, such as hiding or fleeing.

###### 2. PTSD Symptoms

The participants illustrated the profound effect and lasting impact of traumas in their mental health. Through their lived experiences, participants described a range of psychological responses consistent with post-traumatic stress disorder, revealing the complexities of their emotional landscapes. Participants frequently described experiencing intrusive thoughts and flashbacks that transport them back to the traumatic event, causing intense emotional distress. As described by one of the participants:

> *” …whenever I heard gunfire, my heart would race, and I feared that the terrible incident would happen again, … as soon as I walked in, memories of that awful day flooded back, and I found myself crying uncontrollably.” Female, (20’s)*

Additionally sleep disturbances mainly nightmares related to the trauma, were a common symptom as expressed by another participant. Participants also exhibited hyper-vigilance, characterized by an increased state of alertness and constant scanning for potential threats. The emotional toll of PTSD was further illustrated through participants’ descriptions of heightened emotional responses. Many reported the physical symptoms they frequently experiences associated with anxiety which manifest as panic attacks during reminders of the trauma. These intense emotional reactions often led to irritability and anger towards family members, straining relationships. Persistent feelings of sadness and hopelessness also significantly affected the participants daily functioning and overall well-being.

###### 3. Coping Mechanisms

Participants described various coping mechanisms employed to manage the effects of trauma on their lives, revealing both adaptive and maladaptive strategies.

Participants emphasized the importance of seeking emotional support from family and community members during difficult times. Most of the participants also stated that praying provides them comfort and a sense of meaning helping them to cope with emotional pain. In the words of one participant:

> *“I also spend a lot of time praying. It gives me a sense of peace and reminds me that there’s a higher purpose to everything, even when things are so hard.” Female, (30’s)*

Participants reported actively avoiding places or situations that trigger memories of trauma, limiting their daily activities and social interactions due to fear or anxiety associated with those triggers. Some participants also reported that the news about the conflict bring back there trauma and they become extremely stressed so they stated that they prefer to avoid hearing of these Medias. Some participants turned to alcohol or Khat as a means of escape from emotional pain, leading to further complications in their lives and mental health challenges. As described by one of the participants:

> *“… I try to deal with my feelings is by chewing khat. It’s something I started doing more often since the conflict began. In my youth, I drank alcohol sometimes, but I have reduced that a lot over the years. Now, chewing khat helps me forget about the ongoing conflict, even if it’s just for a little while.” Male, (40’s)*

###### 4. Impact on Life

The impact of PTSD on individuals extends into various aspects of their lives, particularly affecting their relationships and daily functioning. The emotional toll of PTSD often leads to increased conflict or emotional distance within families, as participants grapple with unregulated emotions stemming from trauma-related stressors. Participants reported withdrawing from social interactions due to fear leading to isolation during critical times.

Participants noted changes in their interactions with children due to heightened anxiety levels, resulting in increased protective behaviors or overreactions. They also reported a loss of motivation at work due to trauma-related stressors impacting their ability to engage in productive activities. Another participant shared her experience of closing her restaurant due to trauma.

> *“After closing my restaurant, I have been at home for the last year, and this has made my depression worse.” Female, (20’s)*

Impaired concentration was another prevalent issue among participants, as intrusive thoughts often interfered with their ability to focus on tasks.

## Discussion

This study found a 36.2% (95% CI; 32.2%, 40.0%) prevalence of PTSD among residents of Bahir Dar, Ethiopia, indicating a significant mental health concern in this conflict-affected region. This prevalence aligns with rates reported in other conflict zones, such as in Ukraine 37.6% (19), and Syrian Kurdish refugees 35% to 38% (20), implying that prolonged exposure to violence can lead to comparable mental health outcomes across different cultural contexts.

Higher prevalence rates have been reported, such as among Palestinians in the West Bank and Gaza (46.5–49.3%) (21), 53.4% in Nepal (22), 61.2% among Rohingya refugee in Bangladesh (23). In Nigeria, 85.5% (5), along with 40.7% in South Sudan (24) and 58.4% in the Gedeo zone of Ethiopia (1). The variation in findings may rise from differences in study participants and settings. For example, the Rohingya study focused on refugees, while other studies examined internally displaced persons (IDPs) in areas like the Gedeo zone of Ethiopia. In contrast, this study involved community residents. Additionally, non-probability sampling was used in Nigeria, which can introduce bias and limit generalizability.

In contrast, some studies shows lower prevalence such as in Israel Israel (29.8%) (25), Croatia (20.7%) (26), Korea (15%) (27), Myanmar (8.1%) (28), in Libya (12.4%) (29), in Central Sudan (12.3%) (5), Northern Uganda (11.8%) (30). and Kenya (10.6%) (31). This might be due to varying in instruments and cut off points used to measure the prevalence of PTSD, such as International trauma questionnaire (ITQ) in Israel, Clinical assessment of post - traumatic stress disorder (CAPS) in Croatia, Mini-International Neuropsychiatric Interview (MINI) in Uganda and trauma screening questionnaire (ITQ) in Kenya. Another reason could be due to the methods they used to collect the data, for instance in Korea they used self-administered techniques. The other explanation for the discrepancies could be because the current study was conducted after one year the war out broke but the city is still undergoing through conflicts while the other studies for instance the research in Kosovo took place seven years following the war (32), and also the study in Uganda was conducted seven years post-conflict (30).

From the multivariable logistic regression it was found that being female, divorced/widowed marital status, depression, suicidal behavior, stressful life event, poor social support, perceived stress, and problematic substance use were statistically significantly associated with PTSD. The in-depth interview also identified the respondents’ traumatic experiences, PTSD symptoms, coping mechanisms, and impact on their life. Aligning with the existing global literatures (25), (31), (33), (34). This higher exposure of females could be due to combination of biological factors, higher risk for gender based violence and societal expectation that raise their emotional burden. The in depth participants enhance these findings, emphasizing the need for gender sensitive mental health interventions.

The absences of a partner in divorced/widowed individuals can significantly heighten emotional distress and vulnerability, may also be more prone to mental health issues such as depression and anxiety, which can coexist with PTSD and complicate recovery. Studies support the notion that diminished social connections increase susceptibility. Theoretical implications suggest that effective interventions must consider the importance of social support systems in mitigating PTSD risk (34) (31).

The participants also described depressive symptoms including persistent sadness, hopelessness, and emotional numbness which associated with PTSD. This may be because depressive symptoms can amplify the emotional burden of trauma and hinder recovery, making individuals more susceptible to PTSD (35). Another reason may also be that depression can lead to rumination on distressing thoughts, difficulties in regulating emotions, and heightened sensitivity to stress, all of which can exacerbate trauma symptoms (36) (37). This is supported by a study conducted in Sudan (38). This implies that pre-existing depression can worsen trauma responses. Therefore, integrated treatment approaches that address both depression and PTSD simultaneously are essential for improving recovery outcomes.

Furthermore, a history of suicidal behavior was associated with developing PTSD. The possible explanation could be that individuals with PTSD often have a history of trauma exposure, significantly increasing the risk of suicidal thoughts and behaviors(39). Another reason might be because individuals with PTSD may experience hopelessness, guilt, and intrusive thoughts about their trauma (40). This finding supported by previous studies (41) (42). This implies that cognitive distortions may lead to increased feelings of despair and a greater likelihood of considering suicide. Practically, this finding underscores the need for targeted interventions focusing on suicide prevention among individuals diagnosed with PTSD.

Respondents who experienced stressful life events experiences such as violence, bombings, and shootings that shattered their sense of security exhibited PTSD symptoms. One participant described witnessing violence and seeing many bodies on the ground.

> *“We saw people being shot, and everyone was running to hide from the gunfire…“We looked out and saw many bodies lying on the ground; … it was a horrifying sight.” Male, (50’s)*

The likely explanation is that repeated or severe stressful events can overwhelm coping mechanisms, increasing vulnerability to PTSD (43). The cumulative psychological impact of such events often leads to feelings of helplessness, anxiety, and fear, which are core features of PTSD. This finding is consistent with other studies, such as research conducted in Nepal during armed conflict. Theoretically, these results highlight how individual and community-level stressors interact to elevate PTSD risk, as described in the Social Ecological Model. Practically, they underscore the need for targeted stress management and trauma-informed interventions for individuals exposed to multiple or severe stressors in conflict settings (44).

Poor social support was also associated with PTSD and it is also supported by previous studies (33), (45). The possible reason might be that lack of social support can foster feelings of helplessness and hopelessness, which are significant predictors of PTSD (46). Practically, enhancing community support networks is essential; initiatives aimed at fostering social connections can significantly reduce feelings of helplessness associated with poor social support.

Participants in the study also described high perceived stress, which was associated with PTSD. For instance one participant’s experience illustrates how persistent feelings of insecurity and worry can contribute to the development of PTSD, highlighting the emotional burden carried by individuals exposed to ongoing violence.

> *“…This fear didn’t go away quickly. It stayed with me for a long time. I felt insecure and worried all the time… It feels heavy inside you every day.” Male, (50’s)*

This could be because individuals with high perceived stress are more sensitive to traumatic events, leading to exaggerated stress responses and an increased risk which is supported by existing literatures (47), (48). Theoretical implications suggest that managing stress is essential for preventing PTSD, highlighting the need for effective stress management strategies. Practically, implementing resilience-building programs focused on stress reduction techniques is vital for individuals exposed to traumatic events.

Respondents with problematic substance use exhibited higher rates of developing PTSD. It is supported by previous studies done in Australian national survey (49), Iraq sand Afghanistan (50). Some of the participants spoke about turning to substances, such as alcohol and khat, as a way of coping with the overwhelming emotional pain and stress following traumatic events.

This might be because those who are experiencing post traumatic symptoms often use substances as a coping mechanism to alleviate distressing symptoms such as anxiety and intrusive memories. This self-medication can lead to problematic substance use, creating a cycle where substance use temporarily alleviates symptoms but ultimately exacerbates them over time. This is supported by existing studies (51, 52). Theoretical implications suggest integrated treatment approaches addressing both conditions simultaneously.

## Strength and Limitation

A major strength of this study is the use of standardized assessment tools for outcome and independent variables, ensuring consistency and reliability in measurement. Additionally, the integration of qualitative methods provided valuable insights into residents’ lived experiences and helped explore underlying factors contributing to trauma. However, the study is not without limitations; social desirability bias may have influenced participants’ responses to sensitive questions, particularly those related to sexual abuse and substance use, as some individuals may have been hesitant to answer honestly.

## Conclusion

Residents of Bahir Dar, Ethiopia experienced a high prevalence of PTSD, highlighting the significant mental health challenges in this conflict-affected community. Female gender, divorced or widowed status, depressive symptoms, suicidal behavior, poor social support, stressful life events, high perceived stress, and problematic substance use were all significant predictors of PTSD. The narratives of participants further illustrate the profound emotional toll of trauma on daily life and well-being. These findings underscore the importance of integrating mental health screening and support into routine care, with a special focus on vulnerable groups. Future research should explore the impact of targeted interventions and community-based support systems on improving recovery and quality of life for individuals affected by conflict-related trauma.

## Data availability statement

The raw data supporting the conclusion of this article will be made available by authors, without undue reservation.

## Ethics statement

The studies involving humans were approved by Bahir Dar University College of Medicine And Health Science Institutional Review Board with the reference number (IRB/2074/2024). Up on explaining the study’s significance to participants, both verbal and written informed consent was obtained. To maintain confidentiality, potential identifiers were omitted in the questionnaire and in the in depth interview and the collected data were securely stored. The studies were conducted in accordance with the local legislation and institutional requirements. The participants provided their written informed consent to participate in this study.

## Authors’ contributions

Medhanit Tadesse Abebie was responsible for the conceptualization, methodology, investigation, data curation, formal analysis, project administration, and writing of the original draft. Minale Tarek contributed to the conceptualization and methodology, provided supervision throughout the research process, and participated in the review and editing of the manuscript. Melak Menberu Guangul contributed to the conceptualization and methodology, supervised the research, and was involved in reviewing and editing the manuscript.

## Acknowledgement

The author thanks all of the data collectors and participants for their contribution to the success of this study.

## References

1. Madoro D, Kerebih H, Habtamu Y, G/tsadik M, Mokona H, Molla A, et al. Post-traumatic stress disorder and associated factors among internally displaced people in South Ethiopia: a cross-sectional study. Neuropsychiatric disease and treatment. 2020:2317–26.

2. Vahia VN. Diagnostic and statistical manual of mental disorders 5: A quick glance. Indian journal of psychiatry. 2013;55(3):220–3.

3. Atwoli L, Stein DJ, Koenen KC, McLaughlin KA. Epidemiology of posttraumatic stress disorder: prevalence, correlates and consequences. Current opinion in psychiatry. 2015;28(4):307–11.

4. Rathod S, Pinninti N, Irfan M, Gorczynski P, Rathod P, Gega L, Naeem F. Mental health service provision in low-and middle-income countries. Health services insights. 2017;10:1178632917694350.

5. Ng LC, Stevenson A, Kalapurakkel SS, Hanlon C, Seedat S, Harerimana B, et al. National and regional prevalence of posttraumatic stress disorder in sub-Saharan Africa: A systematic review and meta-analysis. PLOS Medicine. 2020;17(5):e1003090.

6. Dams J, Rimane E, Steil R, Renneberg B, Rosner R, König H-H. Health-related quality of life and costs of posttraumatic stress disorder in adolescents and young adults in Germany. Frontiers in psychiatry. 2020;11:697.

7. Bock J-O, Luppa M, Brettschneider C, Riedel-Heller S, Bickel H, Fuchs A, et al. Impact of depression on health care utilization and costs among multimorbid patients–results from the MultiCare cohort study. PloS one. 2014;9(3):e91973.

8. Kienzler H, Sapkota RP. The long-term mental health consequences of torture, loss, and insecurity: a qualitative study among survivors of armed conflict in the Dang District of Nepal. Frontiers in psychiatry. 2020;10:941.

9. Verhey R, Chibanda D, Gibson L, Brakarsh J, Seedat S. Validation of the posttraumatic stress disorder checklist - 5 (PCL-5) in a primary care population with high HIV prevalence in Zimbabwe. BMC Psychiatry. 2018;18(1):109.

10. Teshome AA, Abebe EC, Mengstie MA, Seid MA, Yitbarek GY, Molla YM, et al. Post-traumatic stress disorder and associated factors among adult war survivors in Northwest Ethiopia: Community-based, cross-sectional study. Frontiers in Psychiatry. 2023;14.

11. Gelaye B, Williams MA, Lemma S, Deyessa N, Bahretibeb Y, Shibre T, et al. Validity of the Patient Health Questionnaire-9 for depression screening and diagnosis in East Africa. Psychiatry Res. 2013;210(2):653–61.

12. Storebø OJ, Stoffers-Winterling JM, Völlm BA, Kongerslev MT, Mattivi JT, Jørgensen MS, et al. Psychological therapies for people with borderline personality disorder. Cochrane Database Syst Rev. 2020;5(5):Cd012955.

13. Kocalevent R-D, Berg L, Beutel ME, Hinz A, Zenger M, Härter M, et al. Social support in the general population: standardization of the Oslo social support scale (OSSS-3). BMC Psychology. 2018;6(1):31.

14. Aklog T, Tiruneh G, Tsegay G. Assessment of substance abuse and associated factors among students of debre markos poly technique college in debre markos town, East Gojjam Zone, Amhara Regional State, Ethiopia, 2013. Global journal of medical research. 2013;13(4):1.

15. Teferi KA. Psychoactive substance abuse and intention to stop among students of Mekelle University, Ethiopia. MPH thesis. 2011.

16. Motrico E, Moreno-Küstner B, de Dios Luna J, Torres-González F, King M, Nazareth I, et al. Psychometric properties of the List of Threatening Experiences—LTE and its association with psychosocial factors and mental disorders according to different scoring methods. Journal of affective disorders. 2013;150(3):931–40.

17. Asnakew S, Shumet S, Ginbare W, Legas G, Haile K. Prevalence of post-traumatic stress disorder and associated factors among Koshe landslide survivors, Addis Ababa, Ethiopia: a community-based, cross-sectional study. BMJ open. 2019;9(6):e028550.

18. Bastianon CD, Klein EM, Tibubos AN, Brähler E, Beutel ME, Petrowski K. Perceived Stress Scale (PSS-10) psychometric properties in migrants and native Germans. BMC Psychiatry. 2020;20(1):450.

19. Fel S, Jurek K, Lenart-Kłoś K. Relationship between socio-demographic factors and posttraumatic stress disorder: a cross sectional study among civilian participants’ hostilities in Ukraine. International journal of environmental research and public health. 2022;19(5):2720.

20. Ibrahim H, Hassan CQ. Post-traumatic stress disorder symptoms resulting from torture and other traumatic events among Syrian Kurdish refugees in Kurdistan Region, Iraq. Frontiers in psychology. 2017;8:216128.

21. Canetti D, Galea S, Hall BJ, Johnson RJ, Palmieri PA, Hobfoll SE. Exposure to Prolonged Socio-Political Conflict and the Risk of PTSD and Depression among Palestinians. Psychiatry. 2010;73(3):219–31.

22. Thapa SB, Hauff E. Psychological distress among displaced persons during an armed conflict in Nepal. Social Psychiatry and Psychiatric Epidemiology. 2005;40(8):672–9.

23. Riley A, Akther Y, Noor M, Ali R, Welton-Mitchell C. Systematic human rights violations, traumatic events, daily stressors and mental health of Rohingya refugees in Bangladesh. Conflict and Health. 2020;14(1):60.

24. Ng LC, López B, Pritchard M, Deng D. Posttraumatic stress disorder, trauma, and reconciliation in South Sudan. Social Psychiatry and Psychiatric Epidemiology. 2017;52(6):705–14.

25. Levi-Belz Y, Groweiss Y, Blank C, Neria Y. PTSD, depression, and anxiety after the October 7, 2023 attack in Israel: a nationwide prospective study. EClinicalMedicine. 2024;68.

26. Stevanović A, Frančišković T, Vermetten E. Relationship of early-life trauma, war-related trauma, personality traits, and PTSD symptom severity: a retrospective study on female civilian victims of war. European journal of psychotraumatology. 2016;7(1):30964.

27. Ikin JF, Creamer MC, Sim MR, McKenzie DP. Comorbidity of PTSD and depression in Korean War veterans: Prevalence, predictors, and impairment. Journal of Affective Disorders. 2010;125(1):279–86.

28. Fan X, Ning K, Ma TS, Aung Y, Tun HM, Zaw PPT, et al. Post-traumatic stress, depression, and anxiety during the 2021 Myanmar conflict: a nationwide population-based survey. The Lancet Regional Health-Southeast Asia. 2024;26.

29. Charlson FJ, Steel Z, Degenhardt L, Chey T, Silove D, Marnane C, Whiteford HA. Predicting the Impact of the 2011 Conflict in Libya on Population Mental Health: PTSD and Depression Prevalence and Mental Health Service Requirements. PLOS ONE. 2012;7(7):e40593.

30. Mugisha J, Muyinda H, Wandiembe P, Kinyanda E. Prevalence and factors associated with Posttraumatic Stress Disorder seven years after the conflict in three districts in northern Uganda (The Wayo-Nero Study). BMC Psychiatry. 2015;15(1):170.

31. Jenkins R, Othieno C, Omollo R, Ongeri L, Sifuna P, Mboroki JK, et al. Probable post traumatic stress disorder in Kenya and its associated risk factors: a cross-sectional household survey. International journal of environmental research and public health. 2015;12(10):13494–509.

32. Morina N, Schnyder U, Klaghofer R, Müller J, Martin-Soelch C. Trauma exposure and the mediating role of posttraumatic stress on somatic symptoms in civilian war victims. BMC Psychiatry. 2018;18(1):92.

33. Carmassi C, Dell’Osso L, Manni C, Candini V, Dagani J, Iozzino L, et al. Frequency of trauma exposure and Post-Traumatic Stress Disorder in Italy: analysis from the World Mental Health Survey Initiative. Journal of Psychiatric Research. 2014;59:77–84.

34. Housen T, Lenglet A, Ariti C, Shah S, Shah H, Ara S, et al. Prevalence of anxiety, depression and post-traumatic stress disorder in the Kashmir Valley. BMJ Global Health. 2017;2(4):e000419.

35. Moges S, Molla G, Mekuria K, Molla N, Girmaw F. Posttraumatic Stress Disorder Among Residents of Conflict-Affected Towns. JAMA Network Open. 2023;6(8):e2329156-e.

36. Carmassi C, Cruz-Sanabria F, Gravina D, Violi M, Bonelli C, Dell’Oste V, et al. Exploratory Study on the Associations between Lifetime Post-Traumatic Stress Spectrum, Sleep, and Circadian Rhythm Parameters in Patients with Bipolar Disorder. International Journal of Environmental Research and Public Health. 2023;20(4):3566.

37. Wild J, Smith KV, Thompson E, Béar F, Lommen MJJ, Ehlers A. A prospective study of pre-trauma risk factors for post-traumatic stress disorder and depression. Psychological Medicine. 2016;46(12):2571–82.

38. Mohamed E, Kheir D. Prevalence of Post-Traumatic Stress Disorder and Depression and Associated Factors Among Internally Displaced Persons in Al-Galgala, Sudan. Neuropsychiatric Disease and Treatment. 2024;20:1155–68.

39. Krysinska K, Lester D. Post-traumatic stress disorder and suicide risk: a systematic review. Arch Suicide Res. 2010;14(1):1–23.

40. Fox V, Dalman C, Dal H, Hollander AC, Kirkbride JB, Pitman A. Suicide risk in people with post-traumatic stress disorder: A cohort study of 3.1 million people in Sweden. J Affect Disord. 2021;279:609–16.

41. Holliday R, Borges LM, Stearns-Yoder KA, Hoffberg AS, Brenner LA, Monteith LL. Posttraumatic Stress Disorder, Suicidal Ideation, and Suicidal Self-Directed Violence Among U.S. Military Personnel and Veterans: A Systematic Review of the Literature From 2010 to 2018. Frontiers in Psychology. 2020;11.

42. Akbar R, Arya V, Conroy E, Wilcox HC, Page A. Posttraumatic stress disorder and risk of suicidal behavior: A systematic review and meta-analysis. Suicide and Life-Threatening Behavior. 2023;53(1):163–84.

43. Sareen J. Posttraumatic stress disorder in adults: impact, comorbidity, risk factors, and treatment. Can J Psychiatry. 2014;59(9):460–7.

44. Thapa SB, Hauff E. Psychological distress among displaced persons during an armed conflict in Nepal. Social psychiatry and psychiatric epidemiology. 2005;40:672–9.

45. Veronese G, Pepe A. Sense of Coherence as a Determinant of Psychological Well-Being Across Professional Groups of Aid Workers Exposed to War Trauma. Journal of Interpersonal Violence. 2015;32(13):1899–920.

46. Dinenberg RE, McCaslin SE, Bates MN, Cohen BE. Social support may protect against development of posttraumatic stress disorder: Findings from the Heart and Soul Study. American Journal of Health Promotion. 2014;28(5):294–7.

47. Xue M, Yuan Y, Chen H, Liu Y, Dai M, Sun H, et al. Perceived stress and symptoms of post-traumatic stress disorder in nurses: A moderated mediation model of maladaptive cognitive emotional regulation and psychological capital. Front Psychiatry. 2022;13:902558.

48. Mei S, Liang L, Ren H, Hu Y, Qin Z, Cao R, et al. Association Between Perceived Stress and Post-Traumatic Stress Disorder Among Medical Staff During the COVID-19 Epidemic in Wuhan City. Front Public Health. 2021;9:666460.

49. Katherine L. Mills BHS, Maree Teesson PD, Joanne Ross PD, Lorna Peters PD. Trauma, PTSD, and Substance Use Disorders: Findings From the Australian National Survey of Mental Health and Well-Being. American Journal of Psychiatry. 2006;163(4):652–8.

50. Petrakis IL, Rosenheck R, Desai R. Substance Use Comorbidity among Veterans with Posttraumatic Stress Disorder and Other Psychiatric Illness. The American Journal on Addictions. 2011;20(3):185–9.

51. Leeies M, Pagura J, Sareen J, Bolton JM. The use of alcohol and drugs to self□medicate symptoms of posttraumatic stress disorder. Depression and anxiety. 2010;27(8):731–6.

52. Logrip ML, Zorrilla EP, Koob GF. Stress modulation of drug self-administration: implications for addiction comorbidity with post-traumatic stress disorder. Neuropharmacology. 2012;62(2):552–64.

